# The role of lifestyle in the association of multimorbidity clusters and dementia risk: a large-scale UK Biobank cohort study

**DOI:** 10.64898/2026.07.05.26357302

**Authors:** Tiago Wiesner, Veerle van Gils, Manjae Kwon, Catherine Calvin, Megan Smith, Sarah Bauermeister

## Abstract

**Introduction:** Multimorbidity clusters have been associated with increased dementia risk. While lifestyle factors may modify dementia risk, their role in multimorbidity clusters remains unclear.

**Method:** Data from UK Biobank was used to identify clusters of chronic conditions using latent class analysis, assess their associations with dementia risk using Cox regression, and potential moderating effects of lifestyle factors.

**Results:** We included 465,175 participants (mean age = 56.52 ± 8.01, 53.87 % female). Five clusters were identified and significantly associated with increased dementia risk, with the cardiometabolic (HR = 2.14, *p* < 0.001) and mental health cluster (HR =1.99, *p* < 0.001) exhibiting the highest risk. Only moderate physical activity lowered dementia risk in the pain-dominated multimorbidity cluster (HR = 0.77, *p* = 0.039).

**Discussion:** Lifestyle factors including physical activity may protect against dementia in specific multimorbidity clusters. Future research involving objective and multiple lifestyle measures is needed.

## Introduction

Dementia is a significant public health challenge, ranking globally as the seventh leading cause of death according to the World Health Organisation [1]. As advanced age is strongly associated with dementia risk, aging populations are a major driver of the increasing prevalence of dementia [2]. The number of dementia cases is predicted to rise to over 130 million by 2050, imposing large financial and social burden on the healthcare system and individuals [2,3,4].

In the current aging population worldwide, more than 50% of adults aged 60 or older are presenting with two or more chronic conditions [5]. Crucially, the risk for dementia appears to grow with every chronic condition at older ages [6,7], highlighting the importance of investigating the intersection of multimorbidity and dementia risk. While the number of morbidities are most commonly used to quantify multimorbidity burden [8], they offer limited insight into the nuanced picture of comorbidity patterns and their associated multiplicative risk of dementia. A few studies have recently started to address this limitation by applying a more granular approach, clustering morbidities together and investigating their impact on dementia risk, as summarised in our previous systematic review [9]. Included studies consistently identified clusters of multimorbidity significantly increasing the likelihood of developing dementia. Common clusters included cardiometabolic-related and mental health/neuropsychiatric-related comorbidities. Corroborating this, a systematic review by Busija and colleagues [10] similarly demonstrated that cardiometabolic and mental health conditions were the most consistently replicated profiles of multimorbidity across 51 studies, highlighting the importance of these disease clusters for dementia risk.

However, not many studies have assessed how lifestyle might be involved in the association between multimorbidities and dementia. Recent reports show that up to 45% of dementia cases is likely caused by modifiable risk factors (e.g., physical inactivity, smoking, alcohol consumption, hypertension, and diabetes) [11]. Consistent with this, multidomain lifestyle interventions such as the FINGER trial and the US POINTER Randomized Clinical Trial [12,13] show great potential for reducing the risk of developing dementia. However, not all studies show consistent results [14,15].

Multimorbidity profiles are complex and may vary amongst individuals, increasing the need for more individualised interventions tailored to patients’ specific needs, highlighting the limitations of previous intervention studies. For example, an anti-inflammatory diet has been shown to reduce dementia risk in individuals with specific cardiometabolic diseases [16], evidencing the potential of targeted strategies based on specific comorbidity profiles. However, this effect may differ for individuals within cardiometabolic multimorbidity clusters, mental health-related clusters, or other combinations of conditions, where the most effective lifestyle interventions may vary compared to those for single diseases. In such cases, the interplay between multiple co-existing conditions may reduce the efficacy, limit the feasibility, or render standard lifestyle recommendations insufficient to meaningfully lower dementia risk, reinforcing the need for more personalised and cluster-specific approaches.

To address this research gap, we aim to investigate the potential moderating role of lifestyle factors in the association between multimorbidity clusters and dementia risk. Analyses are performed using data from the UK Biobank, a large-scale prospective cohort. First, we aim to assess whether previously identified multimorbidity clusters can be replicated and whether they show associations with dementia risk. Building on this, the main objective was to examine how specific lifestyle factors (i.e., diet, physical activity, smoking, and alcohol consumption) may moderate these associations.

## Methods

### Study Design and Participants

Participants were selected from the UK Biobank. UK Biobank is a large-scale and detailed prospective study with over 500,000 participants aged 40-69 years recruited between 2006 and 2010, as decribed previously [17]. Study sites involved 22 assessment centres across the United Kingdom (England, Scotland, Wales). Baseline data included clinical measurements, information on genetics, and other risk factors. The sample for the current study consisted of 465,175 participants, after excluding 228 participants with dementia diagnosis (see supplementary material for ICD 10 codes used to define a dementia diagnosis) at or before baseline and 36,769 participants due to incomplete data. The sample was split into a reference group, consisting of participants who have either zero or one chronic condition, and a group of participants who have two or more chronic conditions. Figure 1 shows a detailed overview of the participant selection. The UK Biobank study was approved in 2011 by the NHS Northwest Multi-centre Research Ethics Committee (11/NW/0382) and approval was renewed in 2016 (16/NW/0274) and 2021 (21/NW/0157). All participants provided written informed consent in accordance with the Declaration of Helsinki.

**Figure 1.**
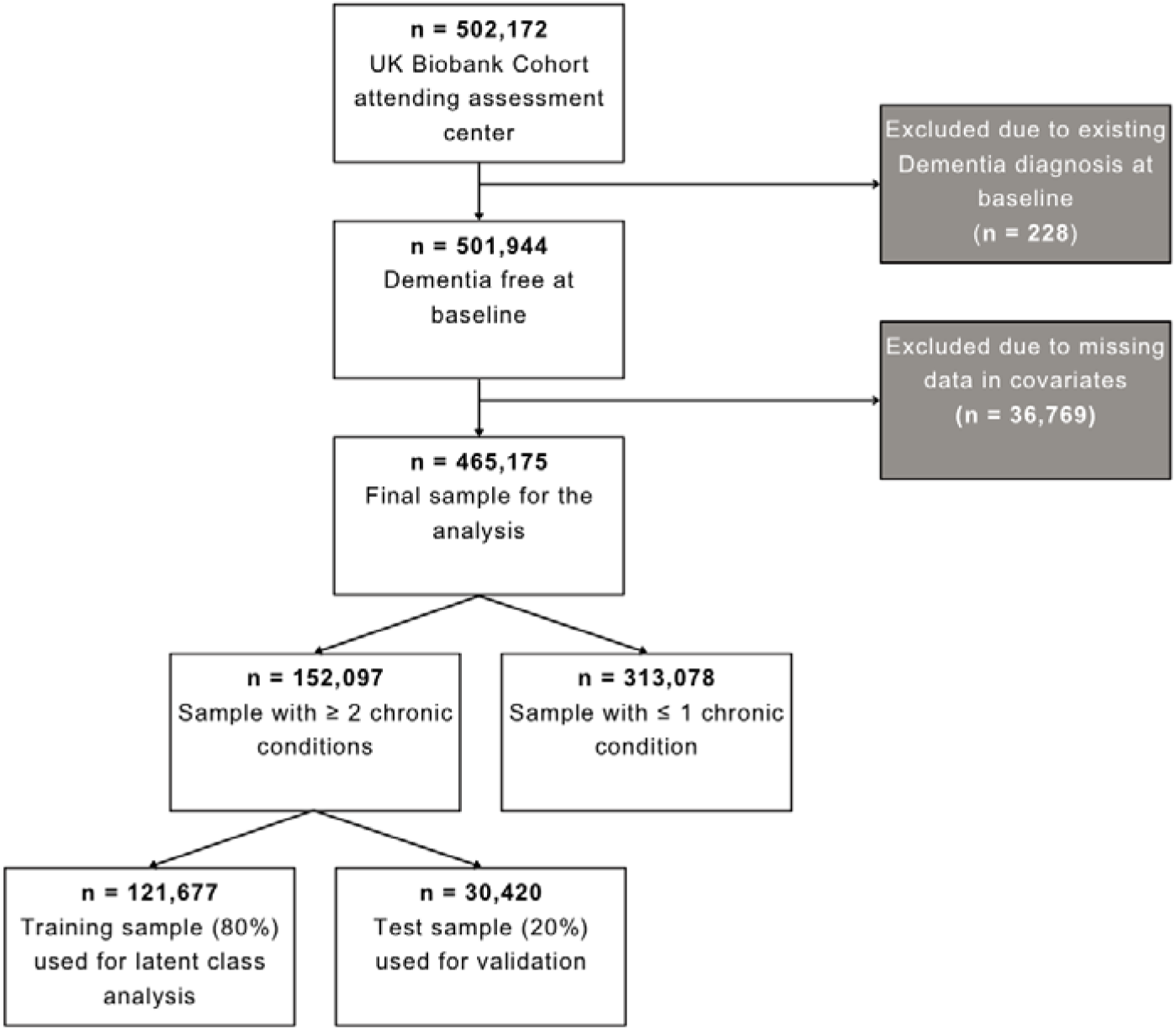
Participant flowchart.

### Multimorbidity and Assessment of Chronic Conditions

We defined multimorbidity as the presence of two or more chronic conditions (see supplementary materials). This is consistent with the methodology for identifying multimorbidity clusters in prior work [18], based on 42 chronic conditions identified by Barnett and colleagues [19]. The assessment of chronic conditions in the UK Biobank is based on self-report at baseline assessment (https://biobank.ctsu.ox.ac.uk/crystal/label.cgi?id=1003).

### Dementia Diagnosis

Dementia was classified according to hospital in-patient records, death registry records, and self-report in the UK Biobank [17]. The ICD-9 and ICD-10 codes used can be found in the supplementary materials.

### Lifestyle measures

Lifestyle factors included physical activity, diet, smoking status, and alcohol consumption and were assessed through a variety of questionnaires. Comprehensive details regarding assessment methods are available in the respective cohort documentation (https://ukbiobank.ac.uk).

Smoking status was dichotomised into current/previous smoker and non-smoker. Diet was dichotomised into participants that consumed less than five portions of vegetable and fruit per day and those that consumed five or more portions of vegetable and fruit per day, aligning with the NHS guidelines [20].

Alcohol consumption was divided into three groups: non-drinkers who consume zero units of alcohol per week, moderate drinkers consuming 1 - ≤13 units per week, and heavy drinkers consuming 14 or more units weekly. This categorisation aims to address potential abstainer bias, occuring when individuals avoid alcohol due to health reasons or because of previous drinking habits [21]. It also reflects evidence suggesting that the risk of dementia increases linearly with the consumption of 14 or more units weekly [22], while no substantial evidence links moderate drinking to an increased risk. This grouping thus aims to capture the J-shaped relationship often observed between alcohol consumption and dementia risk [23].

Physical activity was categorised into three groups of participants who had less than 600 total metabolic equivalent task (MET) minutes per week, 600 - ≤ 3000 MET-minutes per week (with 600 being the recommended minimum) [24], and 3000 or more MET-minutes per week [24]. The MET includes both walking as well as moderate to vigorous physical activity. **Covariates**

Sociodemographic covariates included age in years, sex, education (i.e., college or university, secondary school or vocational qualification, and no qualification), and tertiles of socioeconomic status (based on the Townsend deprivation score) [26], and ethnicity dichotomised into White and non-White [11,27]. In addition, the polygenic risk score (PRS) for Alzheimer’s Disease, split into tertiles [28] as well as C-reactive protein levels, divided into those below 5 mg/L and those at 5 mg/L or above, were included [29].

### Statistical Analysis

#### Latent Class Analysis

A latent class analysis (LCA) was conducted on the training sample (see Figure 1) to extract multimorbidity clusters. As LCA is a probability-based approach, modal assignment (i.e., assigning participants with the highest probability within a cluster) was employed to receive clinically meaningful clusters [18]. This method ensures that all participants are assigned to distinct, non-overlapping clusters, with each patient belonging to only one cluster [30]. Yet, chronic conditions themselves are implicated across clusters with differing probabilities. Assuming a causal relationship between underlying factors and the development of chronic conditions, we expected the multimorbidity profiles of individuals to manifest as discrete clusters. LCA is also a person-centred approach with established use in multimorbidity clustering for grouping individuals with similar profiles [18,31], leading to cluster solutions with improved objectivity, reproducibility, and stability due to its rigorous maximum likelihood estimation. A random training set consisting of 80% of participants within the multimorbidity sample was used to determine the optimal number of clusters (see Figure 1). LCA was applied for 1-10 cluster solutions. We determined the optimal cluster solution by evaluating the sample-size adjusted Bayesian Information Criteria (SABIC), entropy, and expert clinical judgment, while also ensuring that all included clusters represented at least 5% of the training sample. SABIC was chosen as it appears to be among the best fit criteria for identifying the correct number of clusters for categorical LCA models [32]. Entropy quantifies the degree to which participants are distinctly classified into separate clusters. It ranges from zero, indicating substantial overlap in cluster membership probabilities and thus poor separation, to one, indicating that each participant is assigned to a single cluster with near certainty, reflecting optimal separation [18,30]. Higher entropy values, therefore, indicate clearer delineation between clusters and more definitive participant classification. Adequate entropy is essential for the applied modal assignment [33], wherein each participant is allocated to the cluster for which they have the highest membership probability. Lower entropy would increase the likelihood of misclassification, thereby reducing confidence in the reliability of the resulting cluster assignments. In sum, the optimal number of clusters reflects a trade-off between goodness of fit, parsimony, and clinical relevance.

Once the final cluster solution was established, each cluster was characterised by the five most prevalent conditions with an observed-to-expected prevalence ratio greater than one and at a prevalence of at least 5%. This ratio indicates that these conditions occur more frequently within the cluster than would be expected by chance, making them non-random contributors to the cluster’s profile. Finally, to assess the clusters’ validity, the LCA was re-conducted on the test sample, consisting of 20% of the participants, using the best solution from the training set to validate the cluster compositions, sizes, and their associated risk with dementia [18]. The LCA was conducted in R [34]. For the purpose of transparency, the full LCA code and packages used are available from https://github.com/Tiagowi/LCA-multimorbidity-clusters.

### Dementia Risk per Cluster

We examined the association between multimorbidity clusters as predictors and dementia risk as outcome using a Cox proportional hazards (PH) regression model [35]. Hazard ratios (HRs) with 95% confidence intervals (CIs) were obtained. Healthy participants (i.e., <2 chronic conditions) were used as reference and the duration of follow-up as timescale. Follow-up ended based on the latest medical record updates, with censoring dates of October 31, 2022, for England, October 31, 2022, for Scotland, and May 26, 2022, for Wales. Participants who did not develop dementia by the end of the study, withdrew from the study before the end of follow-up, or died, were censored at the time of these events. The Cox PH regression model was adjusted for age, sex, qualification, socioeconomic status, ethnicity, physical activity, diet, smoking status, alcohol consumption, PRS for Alzheimer’s Disease, and baseline inflammation CRP status. To ensure the validity of the model, we assessed multicollinearity among the included covariates using Variance Inflation Factor (VIF), and the PH assumption was assessed by Schoenfeld residuals test and the inspection of log (-log) survival probability plots. Lastly, if dementia cases occurred within one year of baseline [31], they were excluded. Data cleaning and application of Cox PH regression models were conducted in STATA [36].

We extended the analysis by including moderation effects with the included lifestyle factors: physical activity, diet, alcohol consumption, and smoking. This allowed for the investigation of whether and how these lifestyle factors might modify the relationship between multimorbidity clusters and dementia.

### Sensitivity Analyses

Sensitivity analysis was conducted to determine if adhering to more than one healthy lifestyle factor, instead of a single one, would alter the risk of developing dementia. This was explored by examining how combined healthy diet (≥ five portions/day), a physically active life (≥ 600 MET min/week), low to moderate alcohol consumption (0 - ≤13 units), and never being a smoker may reduce dementia risk. In addition, we performed an analysis only including healthy diet and physically active life, given the complex and potentially confounding relationships that alcohol consumption and smoking might have with dementia.

## Results

The final sample included 465,175 participants with a mean age of 56.52 (SD = 8.01) years and 53.87% female. Demographics are shown in Table 1. The training sample used in our main models showed that participants with multimorbidity were generally older, had lower qualification levels, and experienced greater socioeconomic deprivation compared to those without multimorbidity. Additionally, this group was less active, consumed more fruits and vegetables, was more likely to abstain from alcohol, had a lower prevalence of smoking, and exhibited higher CRP levels. Overall, 7,851 participants developed dementia after a mean follow-up time of 13.67 (SD = 1.12) years. In total, 26 participants who developed dementia within one year of baseline were excluded post-hoc.

**Table 1.** Participant characteristics.

| **Participant characteristics** | **All participants**<br>**(n=465,175)** | **No**<br>**multimorbidity**<br>**(n=313,078)** | **Multimorbidity**<br>**(n=121,677)** |
| --- | --- | --- | --- |
| **Age (years)** |  |  |  |
| Mean (SD) | 56.52 (8.01) | 55.27 (8.10) | 59.09 (7.39) |
| **Sex, n(%)** |  |  |  |
| Female | 250,578 (53.87) | 167,145 (53.39) | 66,705 (54.82) |
| Male | 214,597 (46.13) | 145,933 (46.61) | 54, 972 (45.18) |
| **Ethnicity, n(%)** |  |  |  |
| White | 440,790 (94.76) | 296,130 (94.59) | 115,726 (95.11) |
| Non-White | 24,385 (5.24) | 16,948 (5.41) | 5,951(4.89) |
| **Qualification, n(%)** |  |  |  |
| University/College | 153,225 (32.94) | 112,504 (35.93) | 32,715 (26.89) |
| Secondary school<br>& Vocational qual. | 232,307 (49.94) | 157,337 (50.25) | 59,817 (49.16) |
| No Qualification | 79,643 (17.12) | 43,37 (13.81) | 29,145 (23.95) |
| **SES, n(%)** |  |  |  |
| Low deprivation | 158,584 (34.09) | 110,937 (35.43) | 38,094 (31.31) |
| Moderate deprivation | 154,567 (33.23) | 105,574 (33.72) | 39,226 (32.24) |
| High deprivation | 152,024 (32.68) | 96,67(30.84) | 44,357 (36.45) |
| **Physical activity, n(%)** |  |  |  |
| Low | 66,699 (14.34) | 41,856 (13.37) | 19,924 (16.37) |
| Moderate | 184,529 (39.67) | 128,871 (41.16) | 44,530 (36.60) |
| High | 213,947 (45.99) | 142,351 (45.47) | 57,223 (47.03) |
| **Vegetable and fruit consumption, n(%)** |  |  |  |
| < five portions/day | 147,058 (31.61) | 99,577 (31.81) | 38,085 (31.30) |
| ≥ five portions/day | 318,117 (68.39) | 213,501 (68.19) | 83,592 (68.70) |
| **Smoking status, n(%)** |  |  |  |
| Never | 253,711 (54.54) | 179,505 (57.34) | 59,356 (48.78) |
| Previous/current | 211,464 (45.46) | 133,573 (42.66) | 62,321 (51.22) |
| **Alcohol consumption, n(%)** |  |  |  |
| 0 units/week | 121,388 (26.10) | 72,000 (23.00) | 39,141 (32.39) |
| 1 - ≤13 units/week | 145,228 (31.22) | 101,159 (32.31) | 35,223 (28.95) |
| ≥ 14 units/week | 198,559 (42.68) | 139,919 (44.69) | 47,040 (38.66) |
| **PRS Alzheimer's, n(%)** |  |  |  |
| Low | 154,849 (33.29) | 103,866 (33.18) | 40,922 (33.63) |
| Intermediate | 155,087 (33.34) | 104,531 (33.39) | 40,371 (33.18) |
| High | 155,239 (33.37) | 104,681 (33.44) | 40,384 (33.19) |
| **CRP levels, n(%)** |  |  |  |
| Normal | 391,940 (84.26) | 271,289 (86.65) | 96,565 (79.36) |
| Elevated | 73,235 (15.74) | 41,789 (13.35) | 25,112 (20.64) |

Note. SES = socioeconomic status, PRS = polygenic risk score, CRP= C-reactive protein.

### Multimorbidity Clusters

The model with a 5-class solution was considered to be the best model. While the likelihood ratio test remained significant up until the 8-class solution, its tendency to favour more complex models in large samples reinforced our selection of the more interpretable 5-class solution [36]. More information regarding the SABIC and entropy can be found in supplementary materials.

Based on the five most prevalent conditions with an observed-to-expected prevalence ratio greater than one and at a prevalence of at least 5%, the identified clusters were dominated by distinct patterns of multimorbidity. The *cardiometabolic-dominated cluster 1* was characterised by diabetes, hypertension, coronary heart disease (CHD), and stroke, whereas the *cardiometabolic-dominated cluster 2* included hypertension, pain, chronic heart disease (CHD), cancer, and stroke. The *mental health-dominated cluster* was defined by depression, pain, dyspepsia, anxiety, and thyroid disorder, and *the respiratory-dominated cluster* was primarily composed of asthma, psoriasis, and chronic obstructive pulmonary disease (COPD). Finally, a *pain-dominated cluster* emerged, characterised by pain, dyspepsia, cancer, thyroid disorders, and migraine. Figure 2 shows the prevalence of each chronic condition per cluster. All of the five clusters had at least one condition that was present in 100% of individuals. On the foundation of the training sample (n = 121,677), 14,837 (12.19%) of individuals were in the cardiometabolic-dominated cluster 1, 44,005 (36.17%) in the cardiometabolic-dominated cluster 2, 9,991 (8.21%) in the mental health dominated cluster, 24,027 (19.75%) in the respiratory-dominated cluster, and 28,817 (23.68%) in the pain-dominated cluster.

**Figure 2.**
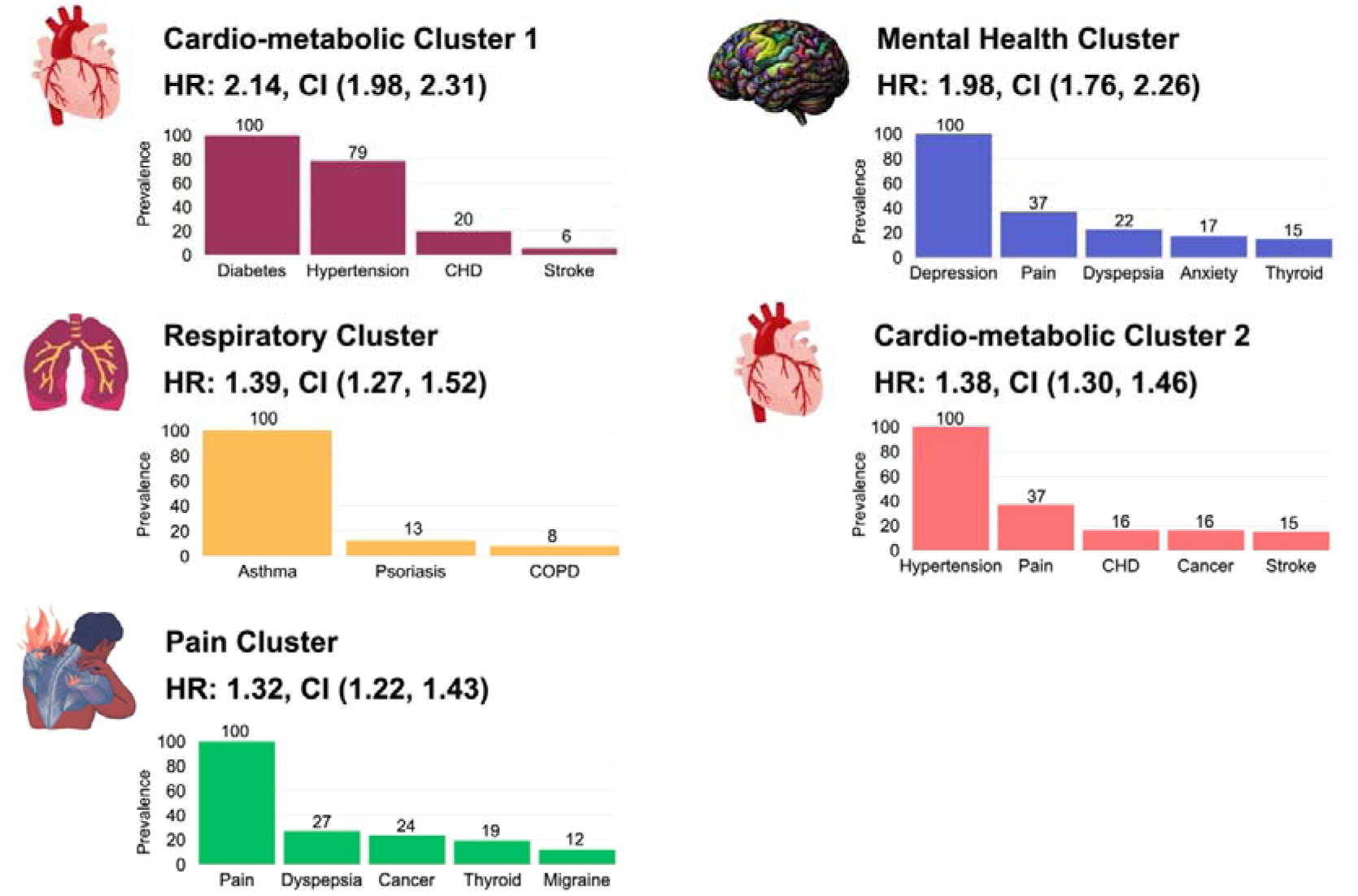
*Prevalence of chronic conditions in each of the multimorbidity clusters.* Only the top five conditions per cluster were included if their observed-to-expected prevalence ratio was greater than one and their prevalence within the cluster was at least 5%. CHD = coronary heart disease, COPD = chronic obstructive pulmonary disease.

### Multimorbidity Clusters and Dementia Risk

As presented in Table 2, the risk of developing dementia was increased in every multimorbidity cluster. The highest risk of developing dementia was observed for the cardiometabolic-dominated cluster 1 (HR = 2.14, 95% CI [1.98 – 2.31], *p* < .001) and the mental health-dominated cluster (HR = 1.99, 95% CI [1.76 – 2.26], *p* < .001). The respiratory-dominated cluster had a HR of 1.39 (95% CI [1.27 – 1.52], *p* < .001), the cardiometabolic-dominated cluster 2 had a HR of 1.38 (95% CI [1.30 – 1.46], *p* < .001), and the pain-dominated cluster had a HR of 1.32 (95% CI [1.22 – 1.54], *p* < .001).

**Table 2.**
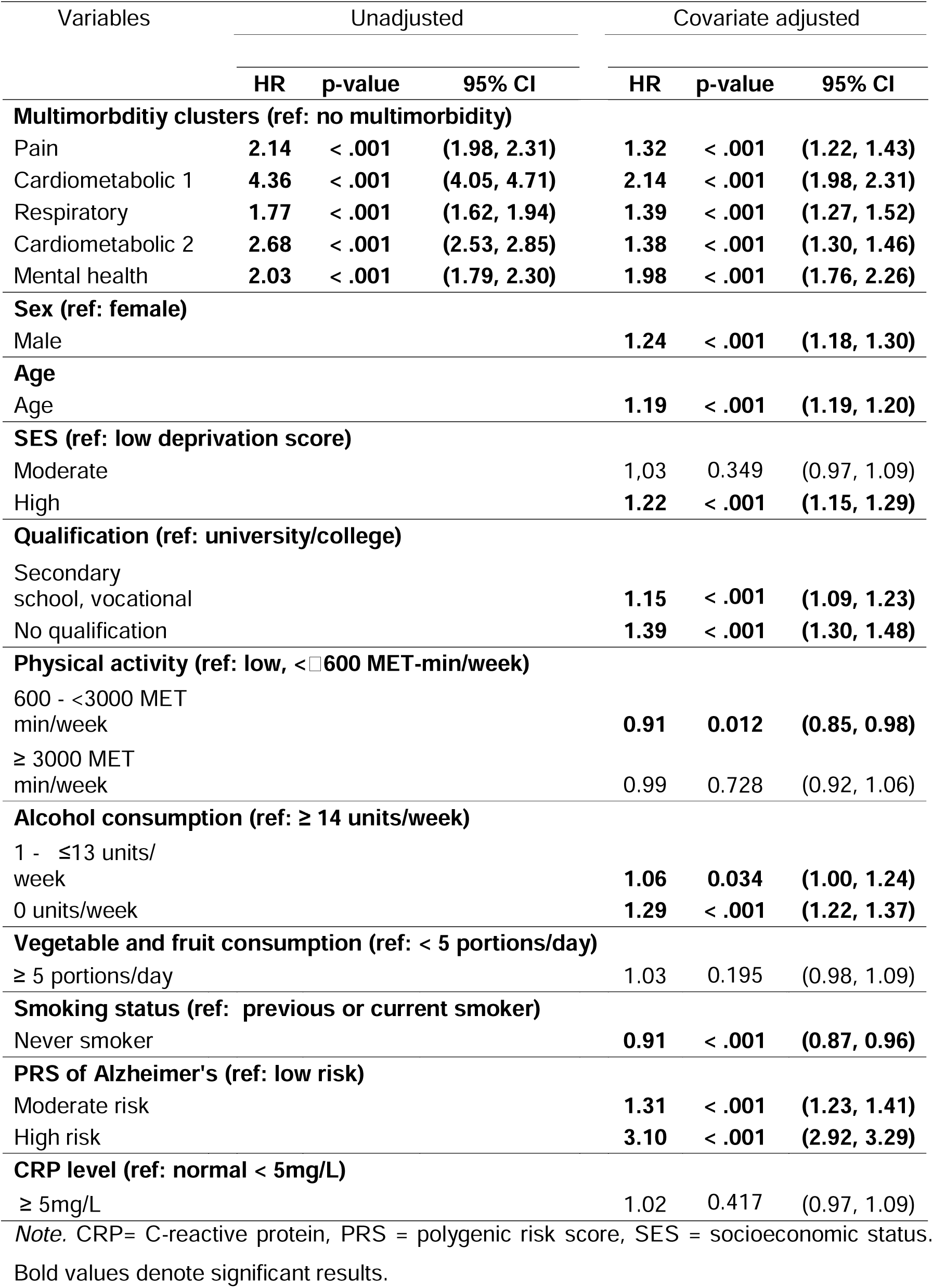
Hazard Ratios (HR) and 95% Confidence Intervals (CI) for Developing Dementia in Unadjusted and Adjusted Cox Regression Model 1.

### Validation

The 5-class solution was re-conducted in the test sample, showing high overlap in cluster composition with the clusters of the main analysis (see supplementary materials). Additionally, the HRs of the clusters show that all clusters were associated with an elevated risk of developing dementia, with the highest risk reported for the cardiometabolic and mental health-dominated clusters, validating the findings from the main model.

### Lifestyle Moderations in Multimorbidity Clusters

The analysis revealed a significant moderation effect between cluster type and physical activity level on dementia risk. The reference group for the multimorbidity cluster analyses comprises individuals without multimorbidity who report an unhealthy lifestyle with low physical activity (<600 MET-min/week), low fruit and vegetable consumption (<5 portions/day), high alcohol intake (≥14 units/week), and a history of current or previous smoking. Among individuals with no multimorbidity, the non-significant HRs indicate no differences in effects on dementia risk for different levels of physical activity (see Figure 3a).

**Figure 3.**
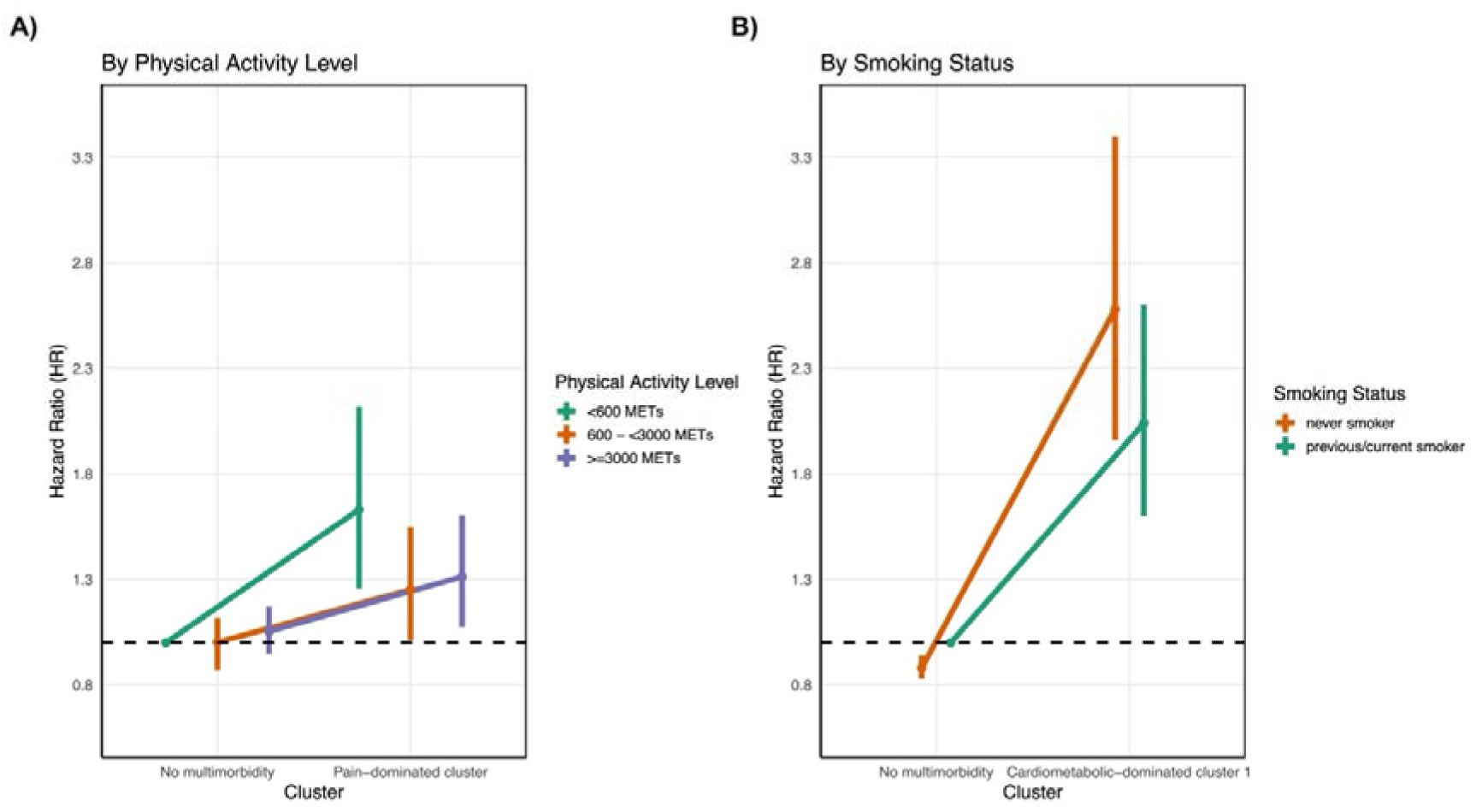
(A) Hazard ratios for dementia risk in the Cardiometabolic-dominated Cluster 1 by smoking status.(B) Hazard ratios for dementia risk in the Pain-dominated Cluster by physical activity level. Vertical lines represent the confidence intervals of the hazard ratios.

Within the pain-dominated multimorbidity cluster, a significant moderation effect for physical activitiy was observed, suggesting that the effect of the pain-dominated cluster on dementia risk differs depending on physical activity level as shown in Figure 3a. Specifically, within the pain-dominated cluster, moderate physical activity was associated with a reduced risk of developing dementia compared to low physical activity (HR = 0.77, 95% CI [0.60 – 0.98], *p* = 0.039). High physical activity also showed a borderline significant moderation with the pain-dominated cluster (HR = 0.8, 95% CI [0.64 – 1.02|, *p* = 0.07), supporting the trend for a possibly protective effect of physical activity for individuals in the pain-dominated cluster. Additionally, a borderline significant moderation was observed between the mental health-dominated cluster and physical activity, with moderate physical activity being associated with a reduced risk of developing dementia compared to low physical activity (HR = 0.70, CI [0.47 – 1.03], *p* = 0.072). Physical activity did not show a moderation effect for any of the other multimorbidity clusters.

A significant moderation was observed between the cardiometabolic-dominated cluster 1 and smoking status as shown in figure 3b. Notably, never smokers indicated a significantly alleviated risk for developing dementia compared to previous and current smokers (HR = 1.27, CI [1.08 – 1.48], *p* = 0.003). Given this controversial finding, an additional analysis of the death rates among clusters for previous and current smokers and non-smokers revealed that smokers in the cardiometabolic-dominated cluster 1 were more likely to die during the follow-up period (approximately 25.6% mortality) compared to other clusters (see Table A4 for more information). This disparity in mortality rates suggests a potential survival bias in the cardiometabolic-dominated cluster 1, whereby a greater proportion of smokers with adverse health outcomes may have died prior to dementia onset. No significant moderation effects were observed for diet or alcohol consumption. See supplementary materials for a comprehensive overview of the moderation terms. Addtionally, the combined influence of these lifestyle factors from our sensitivity analysis also did not show a statistically significant effect (*p* > 0.05).

### Assumption Checks

Results of VIF did not indicate any influential multicollinearities. For the Schoenfeld residual test, the model indicated some statistically significant violations of the PH assumption. However, given the large sample size, some cases may have distorted the meaningfulness of these results [38]. Although minor deviations were noted in the log-log plots (see supplementary materials), the general trends support cautious interpretation of the results without necessitating additional model adjustments.

## Discussion

The present study investigated the association between multimorbidity clusters and dementia risk, and examined potential moderating roles of lifestyle factors within this relationship. Five multimorbidity clusters were identified (cardiometabolic-dominated cluster 1 and 2, mental health-dominated cluster, respiratory-dominated cluster, and pain-dominated cluster), and all were significantly associated with dementia risk, with the highest risk being found for the cardiometabolic-dominated cluster 1 and the mental health-dominated cluster. Moderation effects were observed, indicating that increased physical activity within the pain-dominated cluster was associated with protective effects. Additionally, increased smoking within the cardiometabolic-dominated cluster 1 was linked to a decreased risk of dementia, but might be driven by premature death in smokers. Other lifestyle factors did not show significant moderation effects.

The specific multimorbidity clusters identified in the present study were consistent with previous findings [9]. A cardiometabolic cluster and the mental health cluster showed the highest risk of developing dementia, as consistently indicated by previous studies. Apart from one other study [18], no other study examining the association between multimorbidity clusters and dementia risk has identified a pain-related cluster within the UK Biobank [31,39,40]. This variation underscores differences in the frameworks used to define and categorise chronic conditions across studies [9], and the potential differences in clustering method, which were highlighted by another systematic review [10].

### Influence of lifestyle

It has been repeatedly shown that physical activity is a neuroprotective lifestyle factor reducing dementia risk [40]. Our study indicated that, specifically within the pain-dominated cluster, participants meeting recommended physical activity levels had a lower risk of developing dementia than those who did not. This illustrates the potential benefits of being physically active when experiencing chronic pain with other comorbidities. While conditions such as cancer have shown equivocal associations with dementia [42–44], pain may be a potential driver amplifying dementia risk. Physical activity could play an important role within this cluster, as it has been shown to attenuate pain, reduce stress, and ameliorate depressive symptoms [46–48], all of which are associated with a surge in dementia risk [49–51].

Physical activity has been shown to enhance the production of endogenous opioids, thereby contributing to pain relief [53]. This may be particularly important given a recent study indicating that individuals using opioid analgesics, many of whom present with comorbid conditions, may be at an increased risk of developing dementia [54]. These findings underscore the potential value of adding more cost effective lifestyle-based interventions for pain management, rather then relying solely on pharmacological treatments. Similar positive effects of physical activity may occur in the mental health-dominated cluster, but should be further explored. No effects were observed in cardiometabolic or respiratory clusters. Although speculative, the harmful pathophysiological mechanisms underlying cardiometabolic and respiratory-dominated clusters may be less easily reversible once established.

Other lifestyle factors did not show protective effects. Surprisingly, we found that current or previous smokers showed a lower risk of developing dementia compared to never smokers in the cardiometabolic cluster 1. This counterintuitive finding may be explained by survivor bias [55], caused by smokers being more likely to die before follow-up (see Table A4), as has been shown by previous studies [56,57]. By accounting for this bias (e.g., adjusting for competing risk), future studies may illustrate the potential benefits of abstaining from smoking, given its crucial impact especially regarding to cardiometabolic conditions.

Fruit and vegetable intake showed no significant moderation, despite prior links to lower dementia risk [58,59]. This may reflect limitations of self-reported measures, or the greater importance of broader dietary patterns such as the MIND diet [60]. Recent evidence also suggests anti-inflammatory diets, measured by the Dietary Inflammatory Index [61], which quantifies dietary parameters (e.g., macro- and micronutrients), may reduce dementia risk among those with cardiometabolic conditions [16], thus highlighting the need for more rigorous dietary assessments. Although heavy alcohol use has been linked to increased dementia risk [22], our study found no effects of alcohol use on the association between multimorbidity clusters and dementia. Similar to other lifestyle factors, the complexity of multimorbidity may reflect a more intricate underlying pathophysiology. This could potentially reduce the protective impact of individual lifestyle modifications against dementia risk.

## Limitations

Despite the strength of this large prospective cohort study, which included a wide range of social, lifestyle and biological factors, some limitations in this study need to be addressed. First, participants in the UK Biobank were found to be generally healthier and less diverse than the broader population [62]. They were less likely to smoke, consumed alcohol less frequently, reported a lower number of health conditions, and most were White, potentially limiting the generalisability of the findings. Both chronic conditions and identified multimorbidity clusters may also change over time. As the present analysis is based solely on cluster assignments identified at baseline, individuals may shift between clusters over time [40] influencing our findings. An unique and new aspect of this study is the exploration of lifestyle factors in relation to the association between multimorbidity clusters and dementia risk. Lifestyle factors were however based on categorical and broad measures of the lifestyle factors, making it difficult to detect specific or subtle effects. It is also worth considering that a sensitivity analysis combining adherence to all healthy lifestyle factors assessed in this study did not moderate the asscoation between multimorbidity clusters and dementia risk. This may be explained by the lack of information regarding other lifestyle factors such as quality of sleep and social contacts. Allthough only 7.9% of the sample was excluded due to missing covariate data, they differed in key characteristics such as sex and CRP levels (see supplementary materials). Future studies may, therefore, benefit from applying imputation methods rather than relying solely on complete case analysis to address this issue more comprehensively.

## Conclusion

We investigated the relationship between multimorbidity clusters and dementia risk, and the potential moderating role of lifestyle factors in this association. Consistent with previous findings, multimorbidity clusters were associated with a significantly increased risk of dementia. Among the lifestyle factors, physical activity showed the most promising potential for reducing this risk within specific multimorbidity clusters. To further explore lifestyle in relation to multomorbidity and dementia, research should incorporate more objective measures of, for example, accelometer data for physical activity, and utilise more comprehensive dietary assessments, such as the MIND diet and the DII, to better elucidate their potential impact on dementia prevention within multimorbidity clusters. However, it is important to recognise that the complexity of multimorbidity clusters may require more objective and holistic lifestyle measures and approaches to effectively reduce risk within these clusters. Preventing the onset of multimorbidity altogether may be one of the most critical strategies for mitigating cognitive decline. While research into multimorbidity clusters remains in its early stages, advancing this field is essential, particularly given the high prevalence of multimorbidity in older adults and its strong association with increased dementia risk. As such, targeting multimorbidity remains a key component in efforts to reduce the overall burden of dementia.

## Data Availability

All data produced in the present study are available upon reasonable request to the authors

https://github.com/Tiagowi/LCA-multimorbidity-clusters

## Supplementary Material

**Figure S1.**
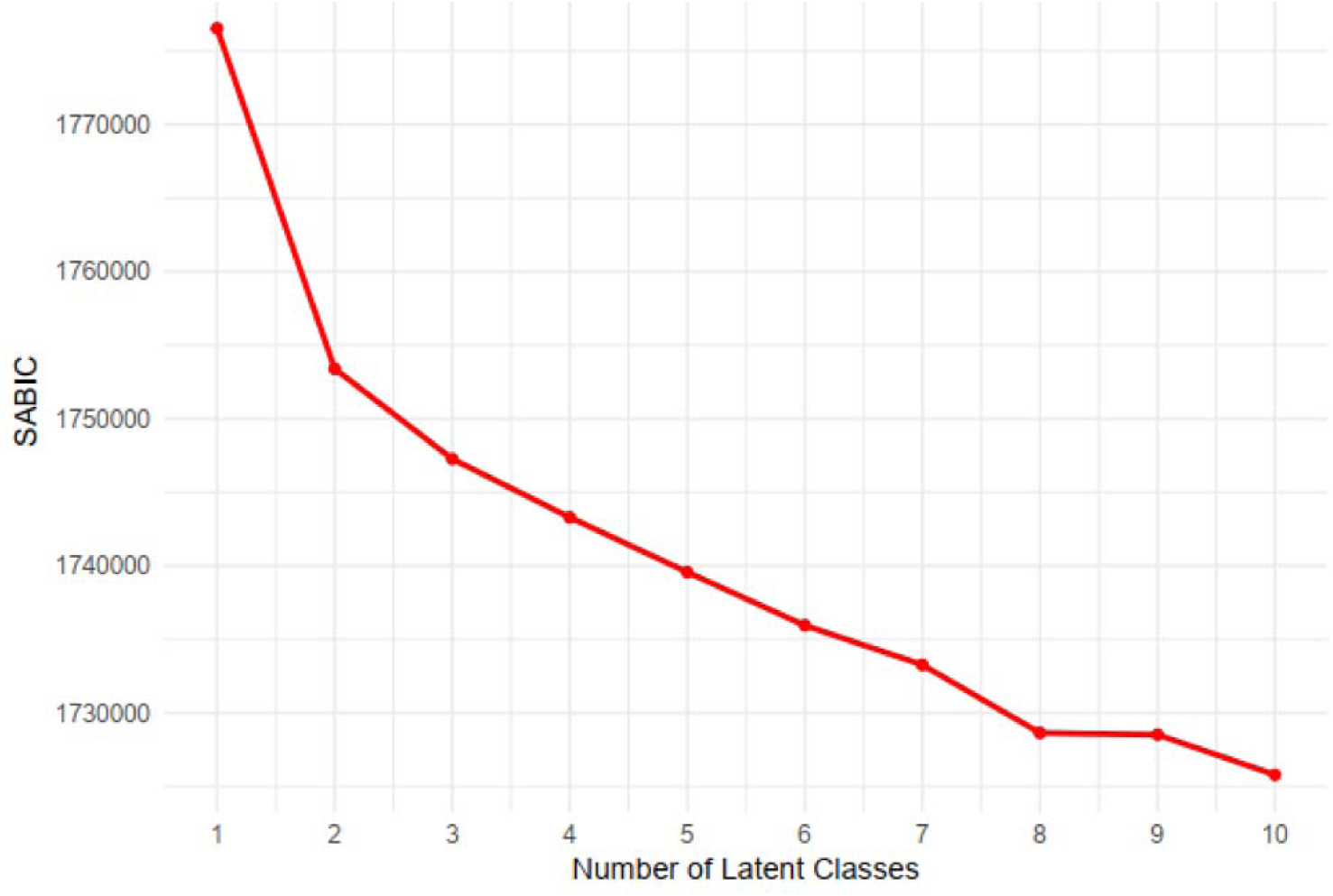
SABIC by number of classes

**Figure S2.**
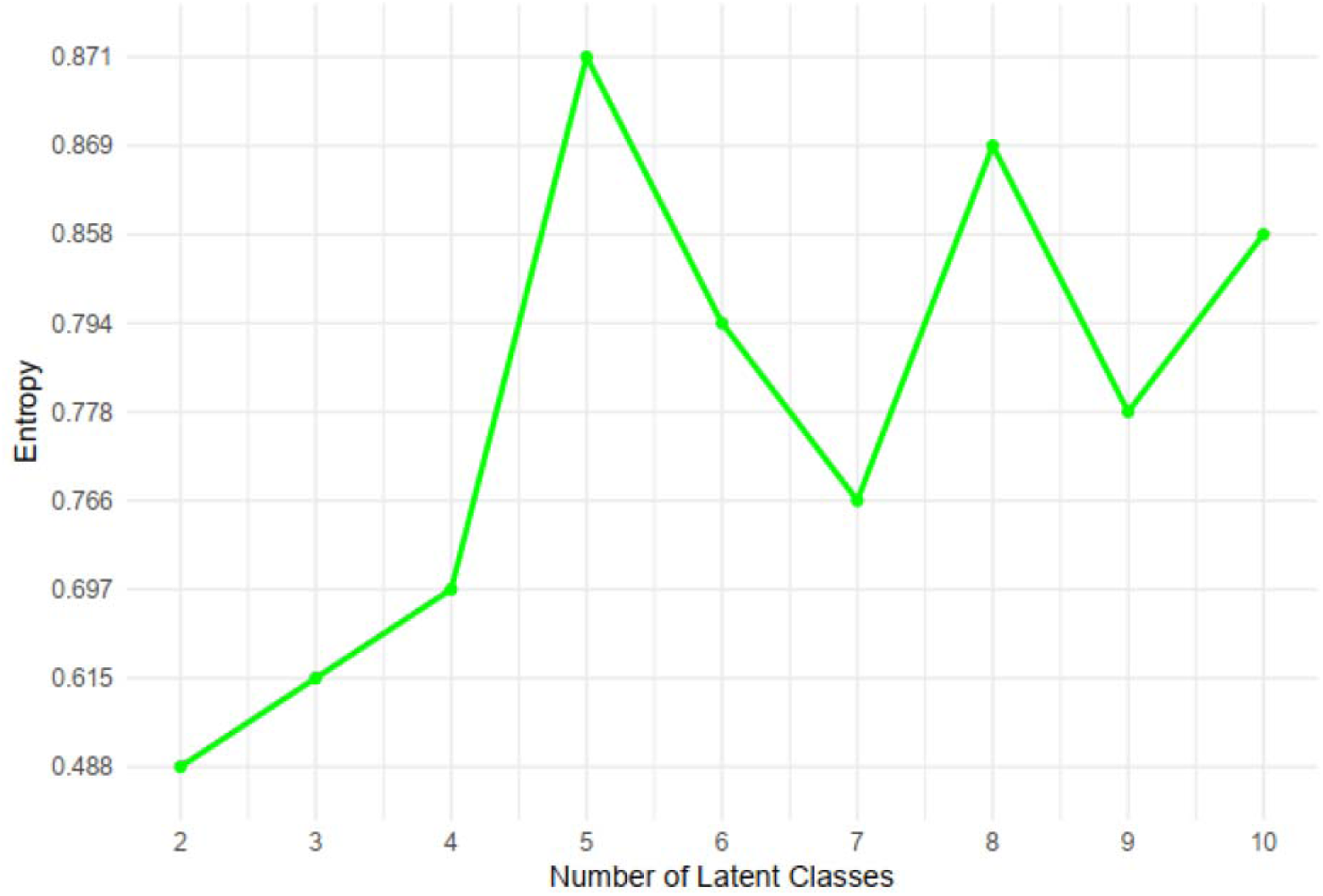
*Entropy by number of classes*

**Figure S3.**
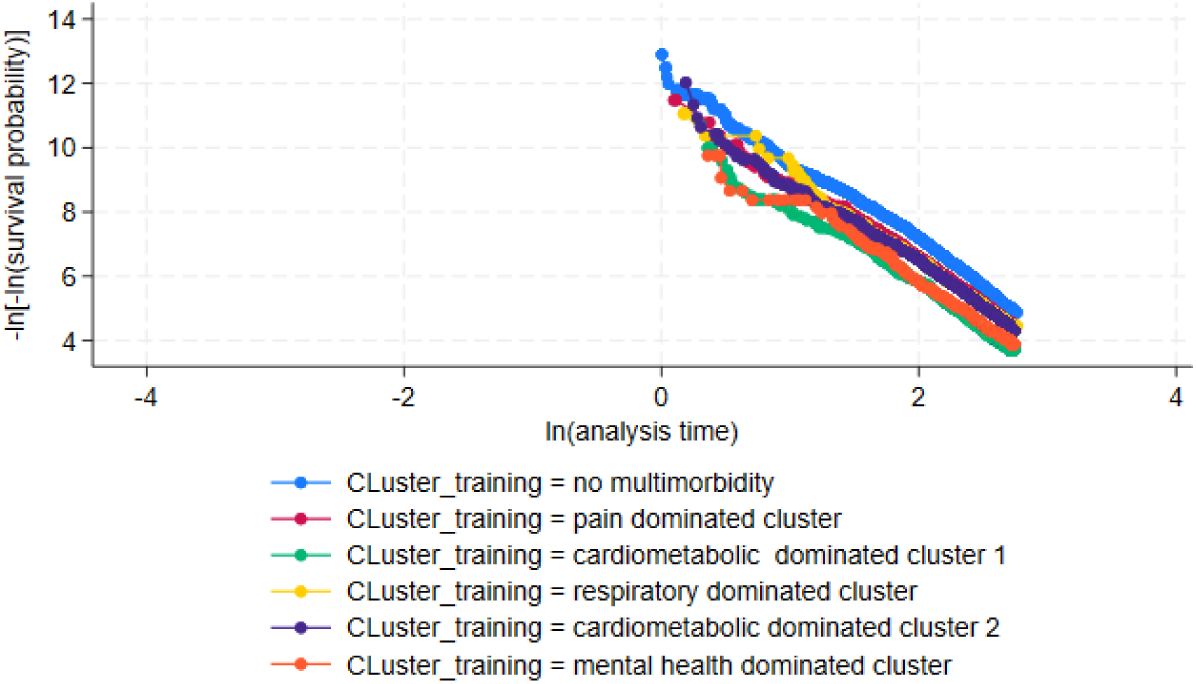
Log (-log) survival probability plots by multimorbidity cluster – adjusted for covariates

**Figure S4.**
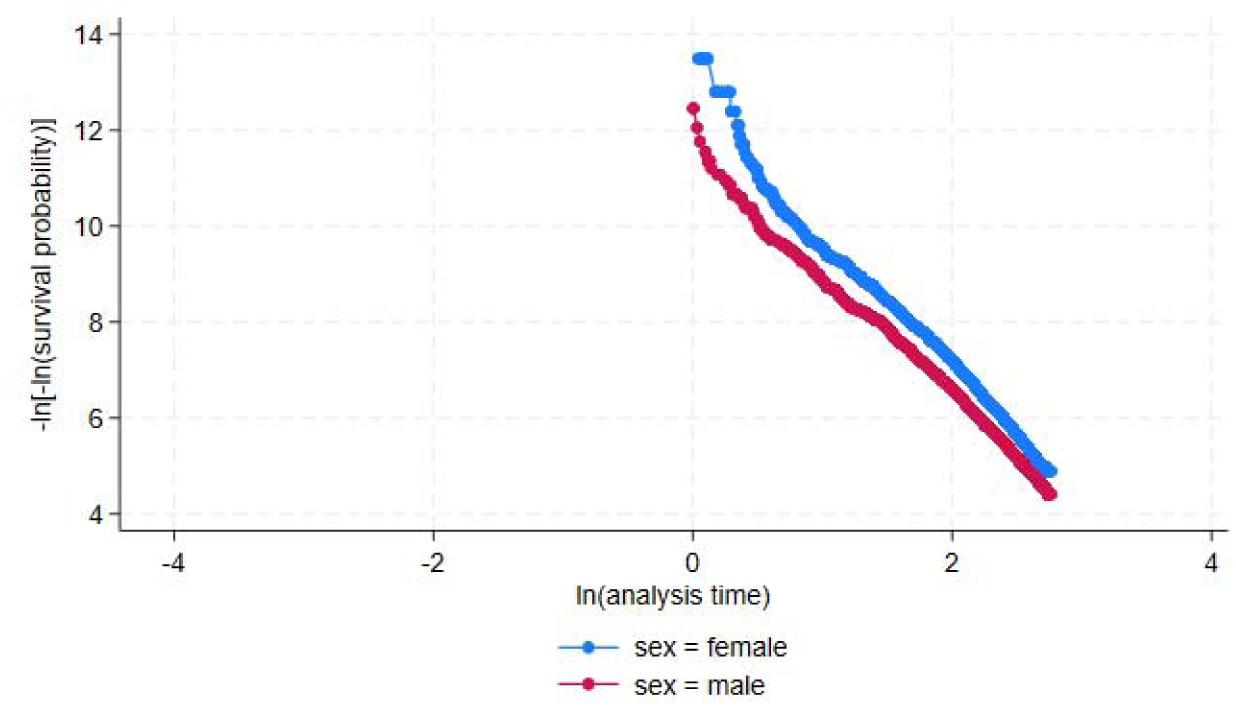
Log (-log) survival probability plots by sex – adjusted for covariates

**Figure S5.**
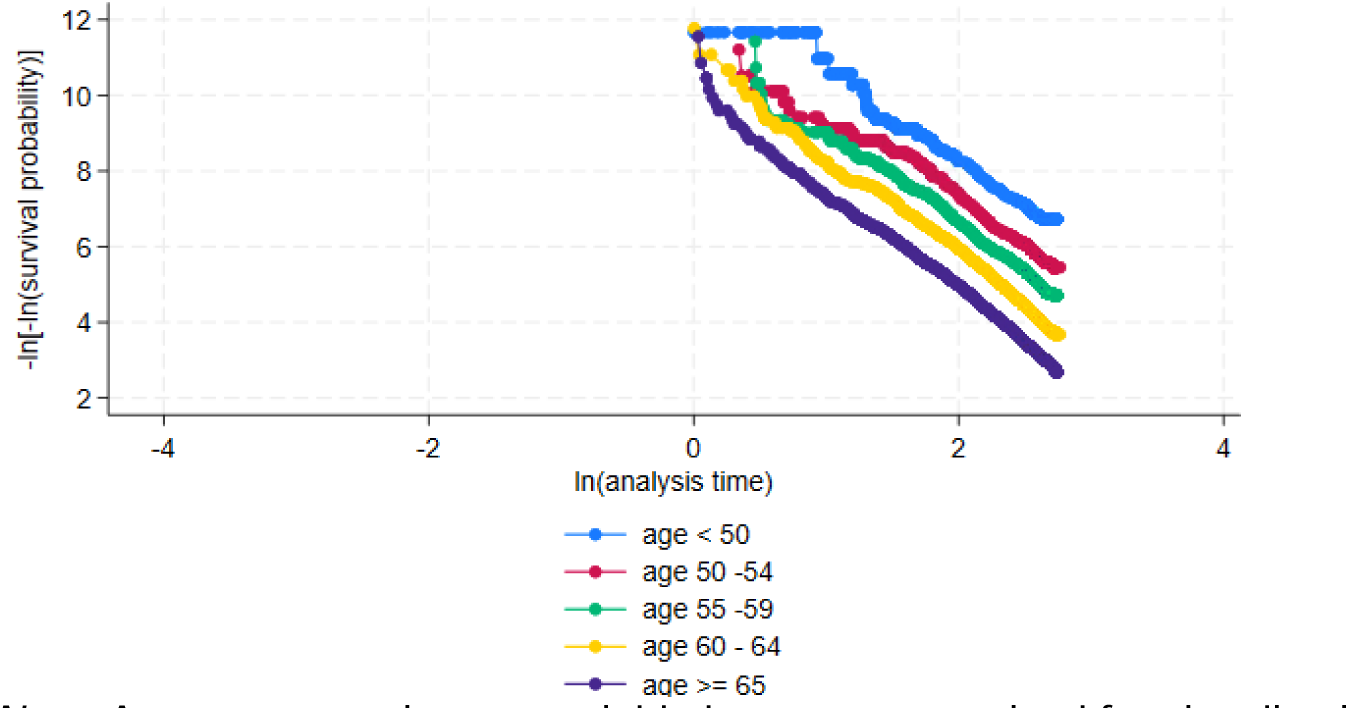
Log (-log) survival probability plots by age – adjusted for covariates Note. Age was a continuous variable but was categorised for visualisation purposes.

**Figure S6.**
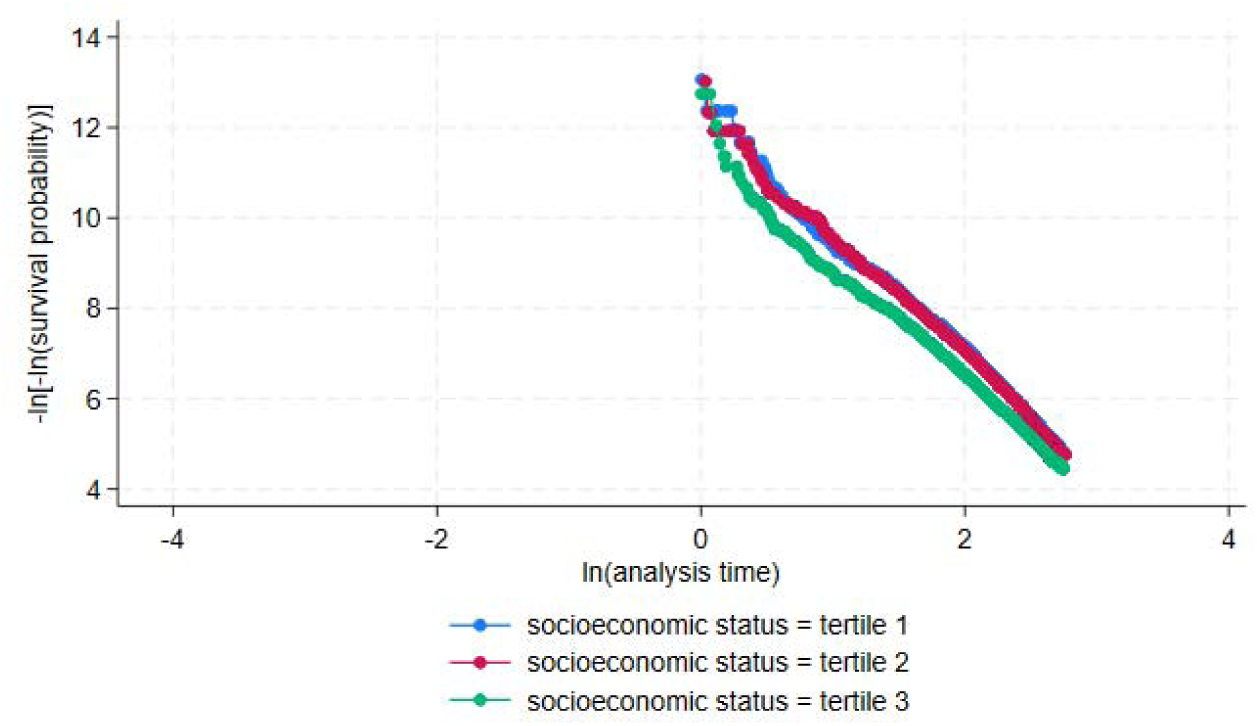
Log (-log) survival probability plots by socioeconomic status – adjusted for covariates

**Figure S7.**
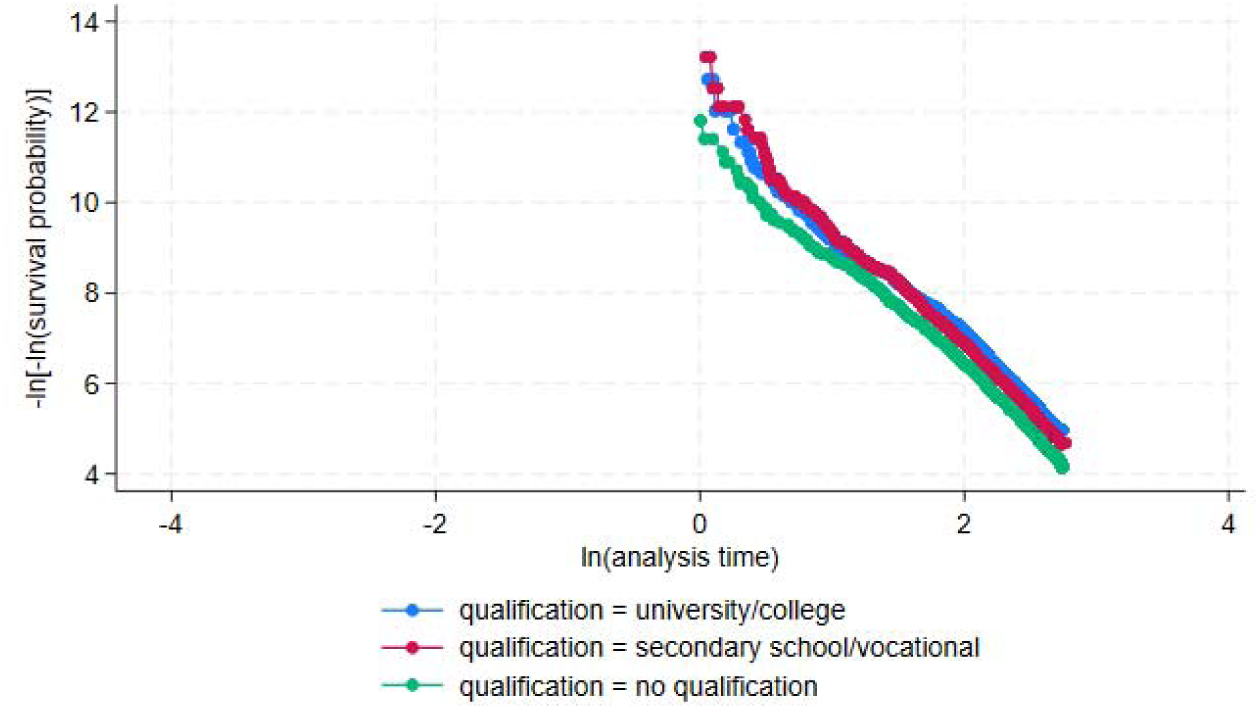
Log (-log) survival probability plots by qualification – adjusted for covariates

**Figure S8.**
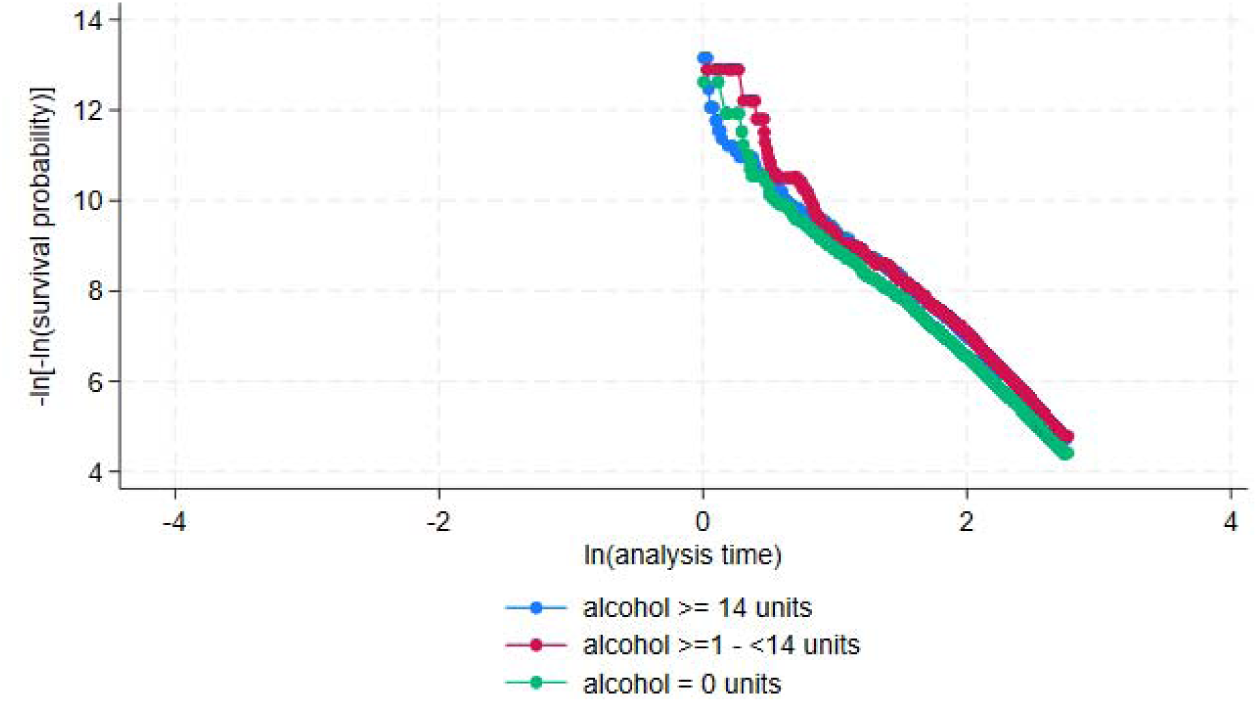
Log (-log) survival probability plots by alcohol consumption – adjusted for covariates

**Figure S9.**
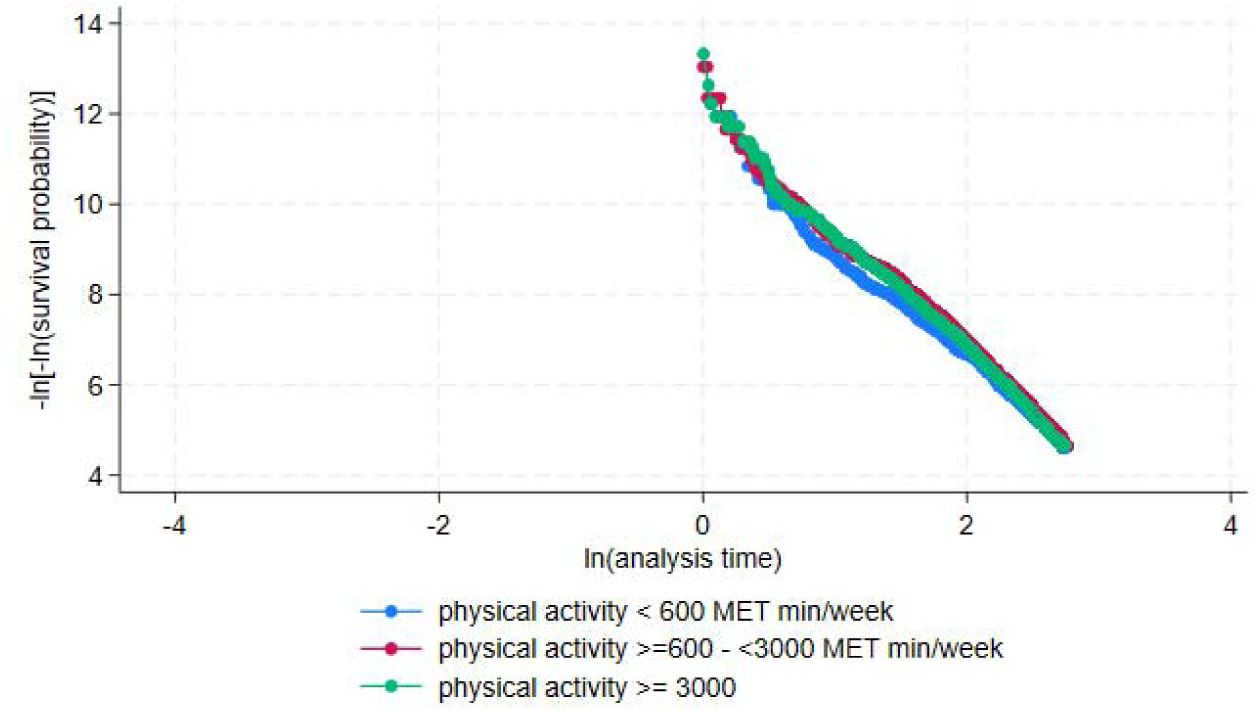
Log (-log) survival probability plots by physical activity – adjusted for covariates

**Figure S10.**
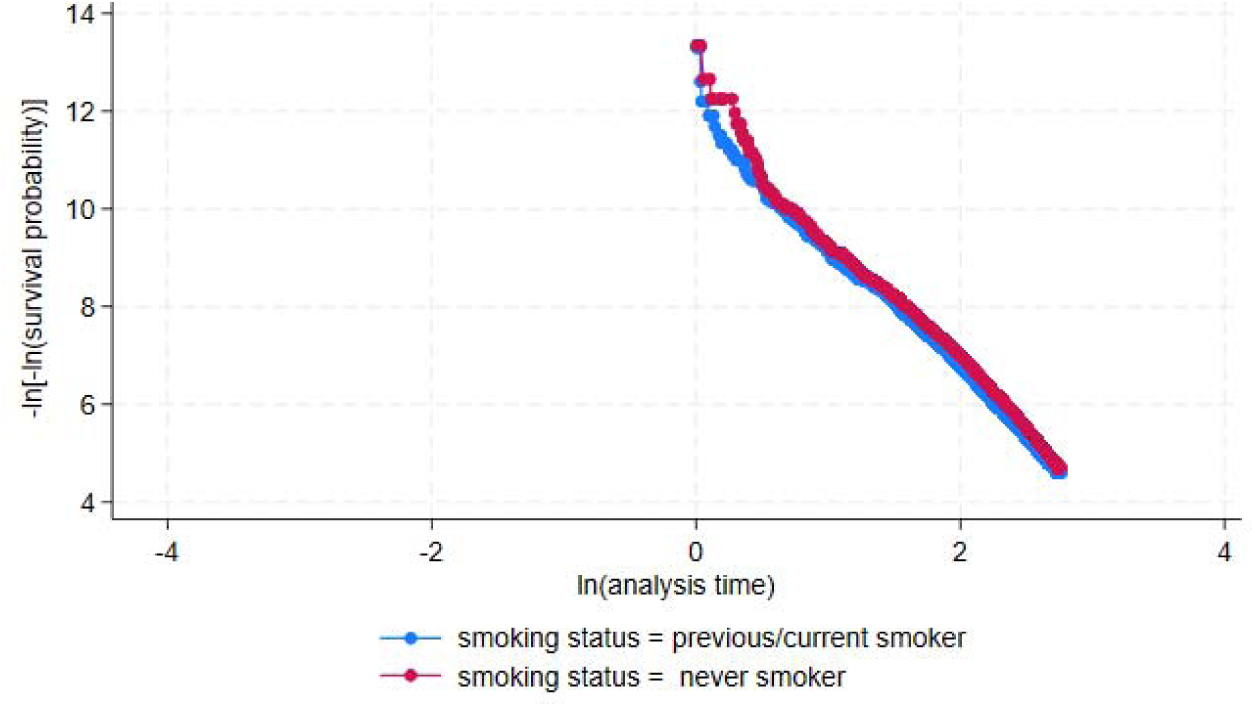
Log (-log) survival probability plots by smoking status– adjusted for covariates

**Figure S10.**
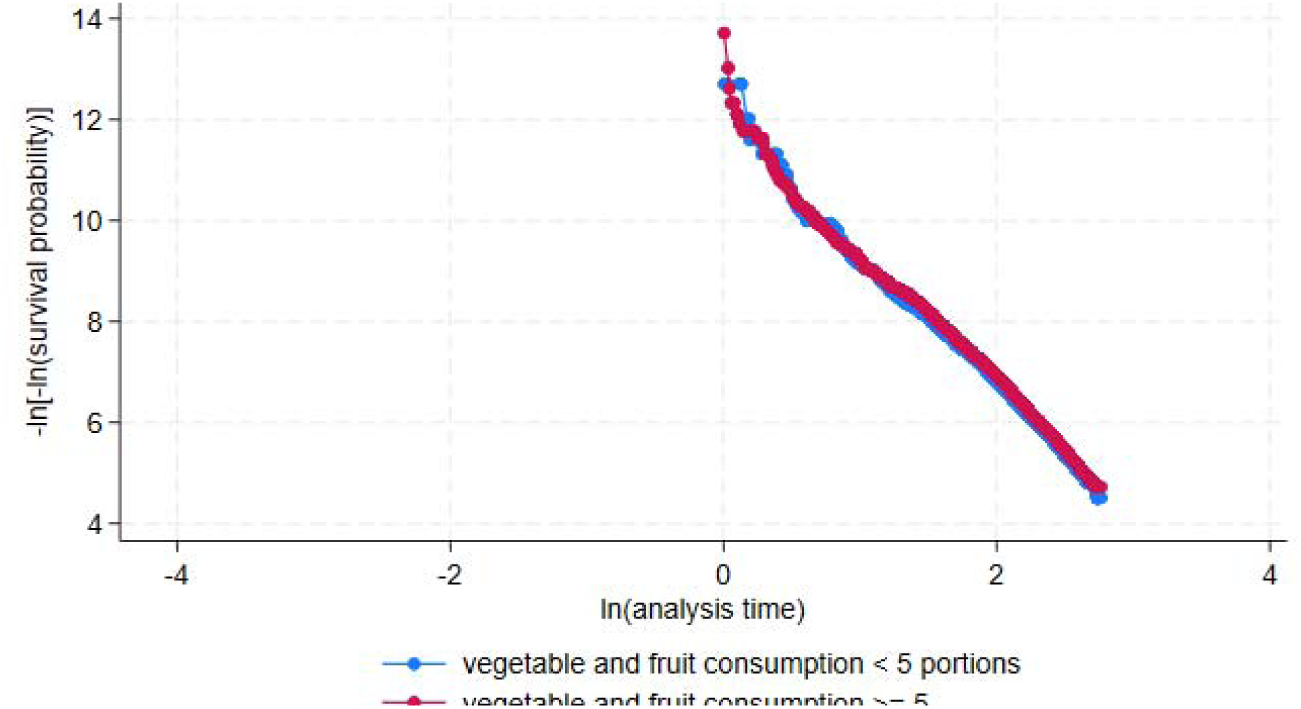
Log (-log) survival probability plots by vegetable and fruit consumption – adjusted for covariates

**Figure S11.**
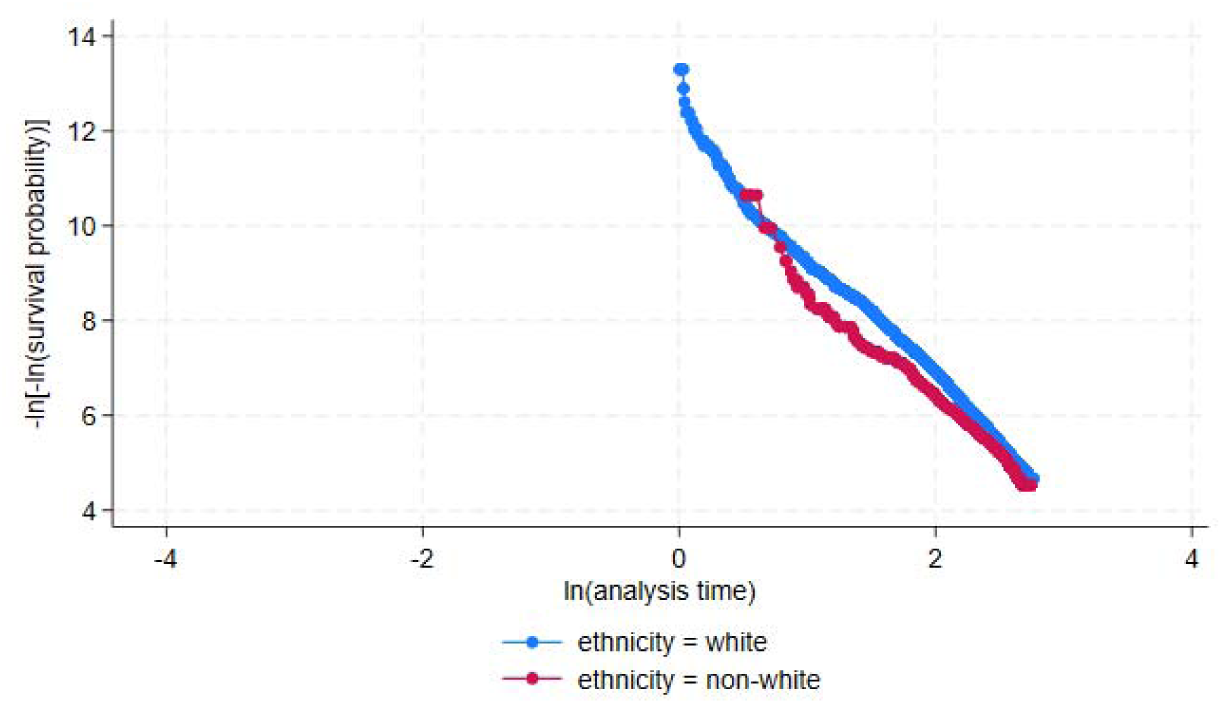
Log (-log) survival probability plots by ethnicity– adjusted for covariates

**Figure S12.**
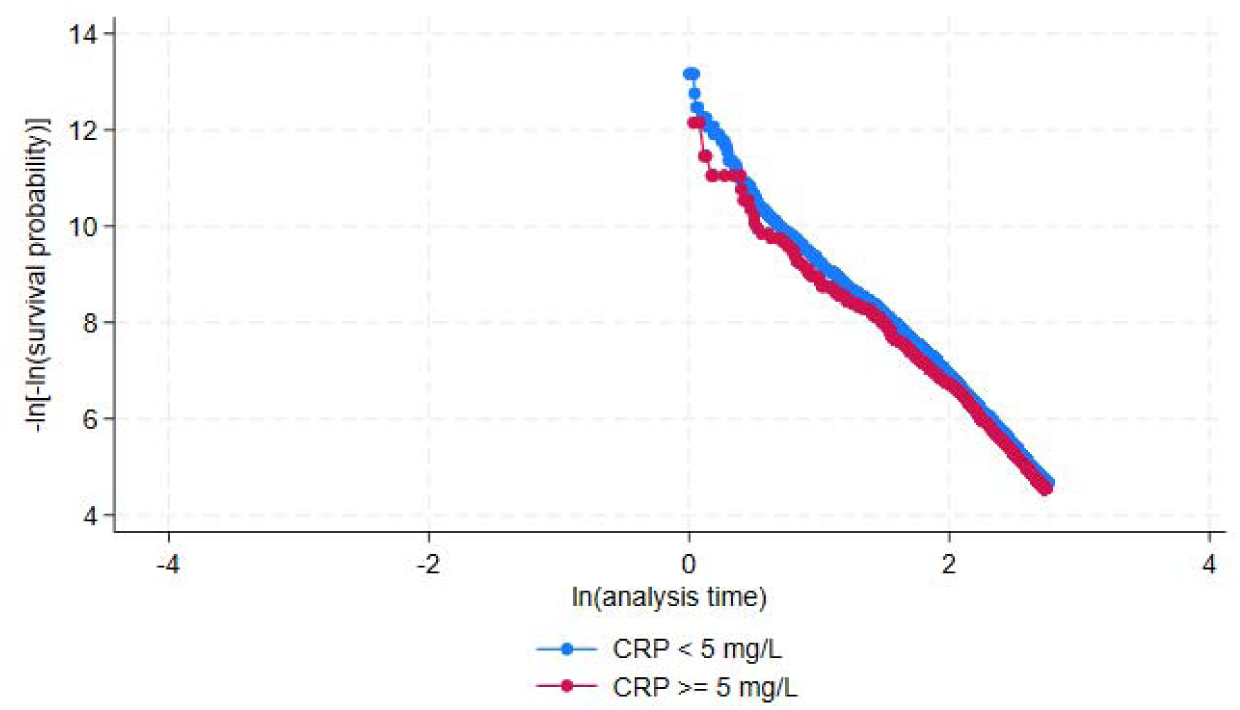
Log (-log) survival probability plots by CRP levels – adjusted for covariates

**Figure S13.**
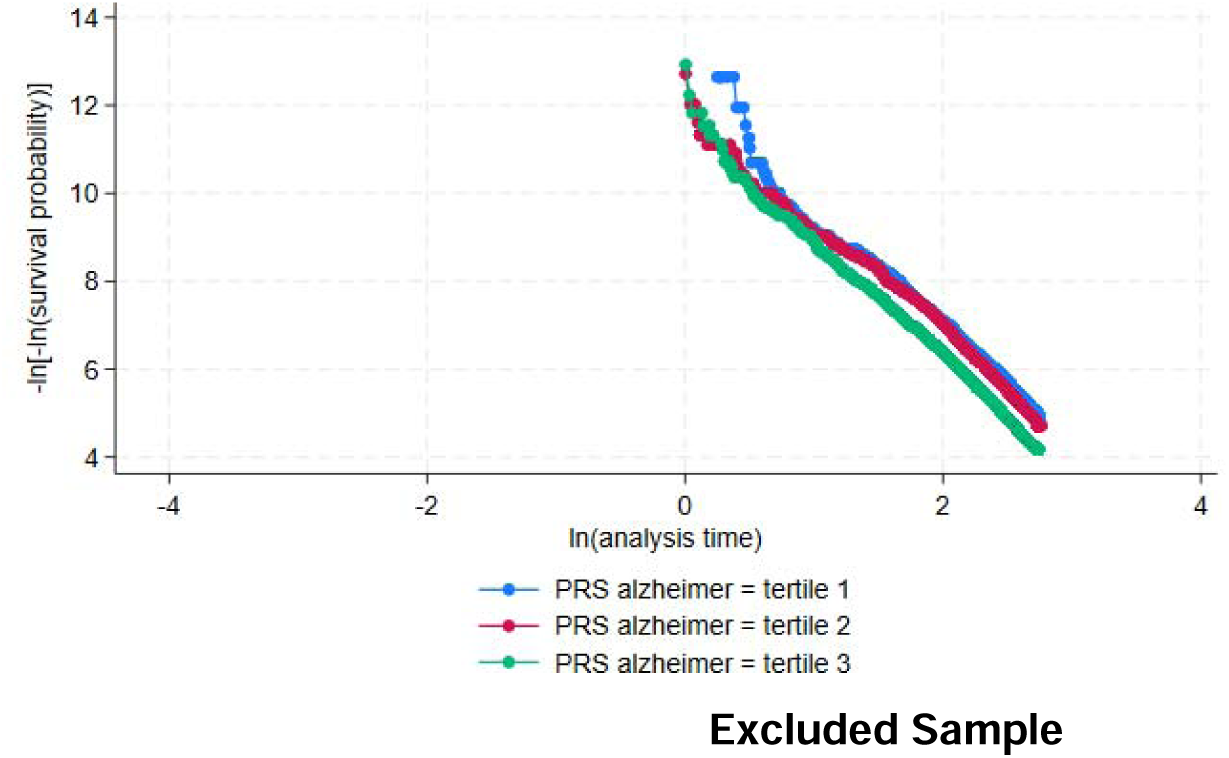
Log (-log) survival probability plots by PRS Alzheimer’s Disease– adjusted for covariates

**Table S1.** Prevalence of the 42 Chronic Conditions Used to Define Multimorbidity.

| Conditions | Prevalence (%) in the Sample |
| --- | --- |
| Hypertension | 122,927 (26.43) |
| Pain | 77,758 (16.72) |
| Asthma | 53,731 (11.55) |
| Dyspepsia | 36,189 (7.78) |
| Cancer | 35,228 (7.57) |
| Thyroid disorders | 26,746 (5.75) |
| Depression | 26,406 (5.68) |
| Diabetes | 22,964 (4.94) |
| Coronary heart disease | 20,774 (4.47) |
| Psoriasis | 16,825 (3.62) |
| Migraine | 13,394 (2.88) |
| Irritable bowel syndrome | 10,967 (2.30) |
| Reumatoid arthritis | 10,128 (2.18) |
| Anxiety | 8,345 (1.79) |
| Stroke & TIA | 8,084 (1.74) |
| Prostate | 7,762 (1.67) |
| Chronic obstructive pulmonary disease | 7,589 (1.63) |
| Osteoporosis | 7,338 (1.58) |
| Intestine disease | 4,959 (1.07) |
| Glaucoma | 4,868 (1.05) |
| Inflammatory bowel disease | 3,944 (0.85) |
| Endometriosis | 3,751 (0.81) |
| Epilepsy | 3,693 (0.79) |
| Atrial fibrillation | 3,403 (0.73) |
| Sinusitis | 2,899 (0.62) |
| Chronic fatigue syndrome | 2,017 (0.43) |
| Schizophrenia or bipolar disorder | 1,814 (0.39) |
| Multiple sclerosis | 1,605 (0.35) |
| Anaemia | 1,384 (0.30) |
| Menieres | 1,289 (0.28) |
| Hepatitis | 1,238 (0.27) |
| Peripheral vascular disease | 1,216 (0.26) |
| Kidney disease | 1,182 (0.25) |
| Bronchiectasis | 1,043 (0.22) |
| **Liver disease** | 904 (0.19) |
| **Parkinson's disease** | 790 (0.17) |
| **Alcohol problems** | 741 (0.16) |
| **Heart failure** | 736 (0.16) |
| **Polycystic ovary** | 571 (0.12) |
| **Anorexia or Bulimia** | 346 (0.07) |
| **Constipation** | 339 (0.07) |
| **Drugabuse** | 84 (0.02) |

**Table S2.** ICD codes used to define dementia diagnosis.

| **ICD version** | **Codes** |
| --- | --- |
| ICD-9 | 331.0, 290.4, 331.1, 290.2, 290.3, 291.2, 294.1, 331.2, 331.5 |
| ICD-10 | F00, F00.0, F00.1, F00.2, F00.9, G30, G30.0, G30.1, G30.8, G30.9, F01, F01.0, F01.1, F01.2, F01.3, F01.8, F01.9, I67.3, F02.0, G31.0, A81.0, F02, F02.1, F02.2, F02.3, F02.4, F02.8, F03, F05.1, F10.6, G31.1, G31.8 |

**Table S3.** Dementia Risk in Test Sample.

| **Multimorbidity clusters** | **Sample (%)** | **Test sample adjusted** |  |
| --- | --- | --- | --- |
|  |  | **HR** | **95% CI** |
| Asthma (100%), Psoriasis (11.6%) and COPD (7.5%) | 6,302 (20.72) | 1.49 | (1.26, 1.76) |
| Depression (100%), Anxiety (15.8%), IBS (9.2%) and migraine (7.6%) | 3,010 (9.89) | 2.09 | (1.68, 2.60) |
| Hypertension (93.7%), Diabetes (36.5%), CHD (27.1%), Stroke (10.2%), Prostate (5.8%) | 9,254 (30.42) | 1.77 | (1.60, 1.96) |
| Cancer (28.5%), Dyspepsia (28.2%), Thyroid (23.8%), Migraine (15.8%) and Psoriasis (12.5%) | 5,017 (16.49) | 1.31 | (1.10, 1.56) |
| Pain (100%), Dyspepsia (19.8%), Migraine (7.1%) and IBS (5.7%) | 6,837 (22.48) | 1.30 | (1.13, 1.50) |

*Note.* The model of the test sample was adjusted for age at baseline, sex, qualification, ethnicity, socioeconomic status, physical activity, diet, smoking status, alcohol consumption, C-reactive protein levels and polygenic risk score of Alzheimer's disease.

**Table S4.** Deaths Among Smokers and Non-Smokers Within Multimorbidity Clusters.

| Cluster | Non-smoker deaths | Previous & current smoker deaths |
| --- | --- | --- |
| No multimorbidity | 7,180 (4%) | 9,768 (7.3%) |
| Pain | 1,083 (7.5%) | 2,015 (14.0%) |
| Cardiometabolic 1 | 1,001 (15.4%) | 2,133 (25.6%) |
| Respiratory | 808 (6.4%) | 1,417 (12.5%) |
| Cardiometabolic 2 | 2,080 (9.9%) | 3,880 (17.0%) |
| Mental health | 283 (5.9%) | 604 (11.7%) |

**Table S5.**

| Clusters by Moderator | Moderator level | HR | P-value | Lower CI | Upper CI |
| --- | --- | --- | --- | --- | --- |
| **Vegetable and fruit consumption** |  |  |  |  |  |
| No multimorbidity | ≥5 portions/day | 1.05 | 0.173 | 0.98 | 1.13 |
| Pain dominated cluster | ≥5 portions/day | 1.05 | 0.602 | 0.88 | 1.24 |
| Cardiometabolic cluster 1 | ≥5 portions/day | 0.98 | 0.856 | 0.83 | 1.17 |
| Respiratory-dominated cluster | ≥5 portions/day | 0.97 | 0.775 | 0.80 | 1.19 |
| Cardiometabolic cluster 2 | ≥5 portions/day | 0.96 | 0.526 | 0.85 | 1.09 |
| Mental health dominated cluster | ≥5 portions/day | 0.82 | 0.110 | 0.62 | 1.05 |
| **Physical activity** |  |  |  |  |  |
| No multimorbidity | 600 - <3000 MET m/w | 1.00 | 0.971 | 0.90 | 1.12 |
| No multimorbidity | ≥3000 MET-m/w | 1.05 | 0.324 | 0.95 | 1.17 |
| Pain dominated cluster | 600 - <3000 MET m/w | **0.77** | **0.037** | **0.60** | **0.98** |
| Pain dominated cluster | ≥3000 MET-m/w | 0.89 | 0.070 | 0.64 | 1.02 |
| Cardiometabolic cluster 1 | 600 - <3000 MET m/w | 0.85 | 0.160 | 0.68 | 1.07 |
| Cardiometabolic cluster 1 | ≥3000 MET-m/w | 0.85 | 0.130 | 0.69 | 1.05 |
| Respiratory dominated cluster | 600 - <3000 MET m/w | 0.79 | 0.110 | 0.58 | 1.06 |
| Respiratory dominated cluster | ≥3000 MET-m/w | 1.05 | 0.714 | 0.80 | 1.38 |
| Cardiometabolic cluster 2 | 600 - <3000 MET m/w | 0.92 | 0.403 | 0.76 | 1.12 |
| Cardiometabolic cluster 2 | ≥3000 MET-m/w | 0.91 | 0.288 | 0.75 | 1.09 |
| Mental health dominated cluster | 600 - <3000 MET m/w | 0.70 | 0.072 | 0.47 | 1.03 |
| Mental health dominated cluster | ≥3000 MET-m/w | 0.92 | 0.625 | 0.65 | 1.30 |
| **Alcohol consumption** |  |  |  |  |  |
| No multimorbidity (reference) | 1 - ≤13 units/w | 1.05 | 0.232 | 0.97 | 1.13 |
| No multimorbidity (reference) | 0 units/w | 1.29 | 0.000 | 1.19 | 1.40 |
| Pain dominated cluster | 1 - ≤13 units/w | 0.96 | 0.680 | 0.79 | 1.17 |
| Pain dominated cluster | 0 units/w | 0.96 | 0.641 | 0.79 | 1.15 |
| Cardiometabolic cluster 1 | 1 - ≤13 units/w | 1.11 | 0.334 | 0.90 | 1.37 |
| Cardiometabolic cluster 1 | 0 units/w | 1.17 | 0.099 | 0.97 | 1.41 |
| Respiratory dominated cluster 1 | 1 - ≤13 units/w | 0.98 | 0.828 | 0.77 | 1.23 |
| Respiratory dominated cluster 1 | 0 units/w | 0.98 | 0.866 | 0.79 | 1.22 |
| Cardiometabolic cluster 2 | 1 - ≤13 units/w | 1.05 | 0.525 | 0.90 | 1.22 |
| Cardiometabolic cluster 2 | 0 units/w | 0.96 | 0.566 | 0.83 | 1.11 |
| Mental health dominated cluster | 1 - ≤13 units/w | 1.25 | 0.194 | 0.89 | 1.75 |
| Mental health dominated cluster | 0 units/w | 0.89 | 0.501 | 0.72 | 1.35 |
| **Smoking status** |  |  |  |  |  |
| No multimorbidity | No smoker | 0.88 | 0.000 | 0.83 | 0.94 |
| Pain dominated cluster | No smoker | 0.98 | 0.761 | 0.83 | 1.14 |
| Cardiometabolic cluster 1 | No smoker | **1.27** | **0.003** | **1.08** | **1.48** |
| Respiratory dominated cluster 1 | No smoker | 0.99 | 0.938 | 0.83 | 1.19 |
| Cardiometabolic cluster 2 | No smoker | 1.05 | 0.436 | 0.93 | 1.19 |
| Mental health dominated cluster | No smoker | 1.21 | 0.141 | 0.94 | 1.57 |

*Note.* m = minutes w = week. Hazard ratios represent differences in effects of multimorbidity clusters by lifestyle factors. The reference group for the multimorbidity cluster analyses comprises individuals without multimorbidity who report an unhealthy lifestyle—specifically, low physical activity (<600 MET-min/week), low fruit and vegetable consumption (<5 portions/day), high alcohol intake ( $\geq 14$ units/week), and a history of current or previous smoking. For analyses focusing on physical activity, comparisons were made against this reference group while varying physical activity levels only, specifically to moderate (600 – <3000 MET-min/week) and high ( $\geq 3000$ MET-min/week) levels, with all other lifestyle factors held constant. A similar approach was applied for dietary behavior, where the level of fruit and vegetable consumption was varied to $\geq 5$ portions/day while other lifestyle variables remained unchanged. The same method was used to examine smoking status, comparing non-smokers to the reference profile while maintaining the same levels of diet, alcohol consumption, and physical activity. Likewise, alcohol consumption was varied to 1 – $\leq 13$ units /week and 0 units/week, respectively, with all other factors held constant to isolate the specific contribution of alcohol intake.

**Table S6.** Excluded Sample.

| Participant characteristics | Analysis Sample (n=465,175) | Excluded sample (n=36,769) |
| --- | --- | --- |
| Age (years) |  |  |
| Mean (SD) | 56.52 (8.01) | 56.65 (8.3) |
| Sex, n (%) |  |  |
| Female | 250,578 (53.87) | 22,501 (61.20) |
| Male | 214,597 (46.13) | 14,268 (38.20) |
| Ethnicity, n (%) |  |  |
| White | 440,790 (94.76) | 31,394 (85.38) |
| BAME | 24,385 (5.24) | 5,375 (14.62) |
| Qualification, n (%) |  |  |
| University/College | 153,225 (32.94) | 7,738 (29.04) |
| Secondary sch& Voc. | 232,307 (49.94) | 13,390 (50.24) |
| No Qualification | 79,643 (17.12) | 5,522 (20.72) |
| SES, n (%) |  |  |
| Tertile 1 | 158,584 (34.09) | 9,614 (26.60) |
| Tertile 2 | 154,567 (33.23) | 11,535 (31.91) |
| Tertile 3 | 152,024 (32.68) | 14,994 (41.49) |
| Physical activity, n (%) |  |  |
| Low | 66,699 (14.34) | 4,203 (11.43) |
| Moderate | 184,529 (39.67) | 10,448 (28.42) |
| High | 213,947 (45.99) | 22,118 (60.15) |
| Vegetable and fruit consumption, n (%) |  |  |
| < five portions | 147,058 (31.61) | 12,820 (34.87) |
| ≥ five portions | 318,117 (68.39) | 23,949 (65.13) |
| Smoking, n (%) |  |  |
| Never | 253,711 (54.54) | 19,527 (57.73) |
| Previous/ current | 211,464 (45.46) | 14,296 (42.27) |
| Alcohol consumption , n (%) |  |  |
| None | 121,388 (26.10) | 8,757 (36.64) |
| Moderate | 145,228 (31.22) | 6,393 (29.09) |
| High | 198,559 (42.68) | 8207 (34.33) |
| PRS Alzheimer's , n (%) |  |  |
| Low | 154,849 (33.29) | 7,083 (34.46) |
| Intermediate | 155,087 (33.34) | 6,818 (33.17) |
| High | 155,239 (33.37) | 6,652 (33.37) |
| CRP, n (%) |  |  |
| Normal | 391,940 (84.26) | 21,535 (58.57) |
| Elevated | 73,235 (15.74) | 15,234 (41.43) |

